# Evaluating human rod photoreceptor function using pixelwise intensity-based optoretinography

**DOI:** 10.1101/2025.09.03.25334736

**Authors:** Gao Yang, Mina Gaffney, Robert F Cooper

## Abstract

More than 50 inherited retinal diseases are known, with rod photoreceptors serving as early indicators in nearly half of them. Given that current clinical imaging modalities cannot resolve the loss of individual rods, rod optoretinograms hold unique promise for transforming the early detection and precise monitoring of retinal disease. They provide a powerful means of detecting early functional changes in diseases marked by rod degeneration, such as Retinitis Pigmentosa, conditions like Age-related Macular Degeneration and certain forms of night blindness, where rod mosaics remain structurally intact, but physiologically impaired. The “optoretinogram” is a relatively new assay that operates through detection of optical changes in cells in response to stimuli. This tool has excellent potential for providing insights into the earliest functional changes of individual photoreceptors, with the potential to assist in the early detection, monitoring, and treatment of retinal diseases.

In this work, we obtained intensity-based optoretinograms (iORGs) from rod photoreceptors using an adaptive optics scanning laser ophthalmoscope. We explore the necessity of both individual rod identification for extracting these waveforms and discuss rod iORG RMS morphology in the context of previously reported cone iORGs.

We found that human rod iORG RMS waveforms have slower implicit times and lower amplitudes than cone iORG RMS waveforms. Additionally, we determined that we obtain very similar iORG RMS metrics using either only rod locations or all pixels where rods reside. The ability to obtain rod optoretinograms without counting individual rods greatly simplifies the functional evaluation of rods and makes the approach more practical and scalable for larger populations and diseased retina.

## Introduction

The human retina contains two types of photoreceptors: cones and rods. Cones mediate high acuity and color vision in well-lit conditions, while rods mediate motion detection and vision in low-light conditions.^1,2^ Photoreceptors may become damaged for various reasons, including metabolic abnormalities or genetic predispositions, resulting in retinal diseases. Specifically, more than 50 documented inherited retinal diseases (IRDs) exist^3-5^; of these, rod photoreceptors are harbingers of early disease in approximately 50% of IRDs and directly affect rod function in IRDs such as Retinitis Pigmentosa,^4,6,7^ and Age-related Macular Degeneration.^5^ Damage to photoreceptors in IRDs can cause visual changes that manifest as night blindness and peripheral vision loss several years before the onset of noticeable disease, restricting activities and negatively impacting the mental and emotional well-being of those affected.^4,5,8^

To evaluate retinal diseases, most functional imaging modalities used in a typical clinical setting, such as multifocal electroretinograms (mfERGs), microperimetry, or dark adaptometry, pool information over a relatively large retinal area and lack sufficient resolution to discern highly localized changes in photoreceptors.^7^ This limitation may lead to an underestimation of the severity of retinal disease, hindering earlier detection and clinical intervention. In contrast, adaptive optics (AO) imaging technology enables high-resolution assessment of the retina at a cellular level. AO-enhanced ophthalmoscopes have been utilized to analyze the structure of the photoreceptor mosaic in health and disease.^8,9^ Over the past decade, there has been a marked increase in the use of these devices for functional assessment, culminating in AO-enhanced assessments of visual acuity, densitometry, and stimulus-evoked changes in photoreceptor morphology. These stimulus-evoked changes, known as optoretinography,^10-29^ enable non-invasive measurement of the function of individual photoreceptors. The “optoretinogram” (ORG) has excellent potential for providing insights into the earliest functional changes of individual photoreceptors, with the potential to assist in the early detection, monitoring, and treatment of retinal diseases.

ORGs are by nature non-invasive and can be obtained *in vivo* by analyzing light in two different ways: by measuring the intensity of the light, termed “intensity-based ORGs” (iORGs), or by measuring the phase change of the light, termed “phase-based ORGs” (pORGs). There have been numerous reports of iORGs^11-13,28-39^ and pORGs ^10,14,16,18,25,26,40-50^ obtained in humans with normal vision, and some reports assessing patients with retinal disease ^13,18,28,29,38^.

Of the three studies that report rod ORGs, one study obtained rod pORGs from two individuals using AO-OCT,^43^ and the others were obtained from single subjects using FF-OCT^41,47^ via segmentation and modeling of rod ORG temporal dynamics, respectively. However, there have been no reports of iORGs from individual rod photoreceptors. As the *en-face* imaging devices that obtain iORGs are less complex and costly to build and maintain than AO-OCTs, there is a substantial benefit to developing the rod iORG on these devices.

To address this gap, we have built upon the iORG acquisition methodology developed by our group^12,31^ with the goal of quantifying rod photoreceptor function in the living human retina. Further, we will determine if adaptive optics laser scanning ophthalmoscope (AOSLO) can capture iORGs from individual rod photoreceptors, and if visualizing every rod photoreceptor is necessary to evaluate rod photoreceptor kinetics.

## Methods

### Research Participants

This study was reviewed and approved by the Institutional Review Board at the Medical College of Wisconsin (PRO#: 36690) and was conducted in accordance with the tenets of the Declaration of Helsinki. Four subjects without known retinal pathology were recruited for this study (**Table 1**). All subjects made a single visit to both the Medical College of Wisconsin and Marquette University to have their randomly selected study eye imaged. All participant IDs are not disclosed to any individuals outside of the research group.

**Table 1.** Participant Demographic Information.

| Subject | Eye | Refraction | Axial Length (mm) |
| --- | --- | --- | --- |
| 36828 | OD | -3.0D+0.25x110° | 24.46 |
| 43556 | OD | -1.5D | 24.30 |
| 86040 | OD | -1.25D+0.50x025° | 23.57 |
| 88375 | OS | -0.25D+1.0x080° | 23.67 |

### Advanced Optics Scanning Laser Ophthalmoscope (AOSLO) System

As previously described,^13^ this study used a customized Apaeros Advanced Optics Scanning Laser Ophthalmoscope (AOSLO) originally constructed by Boston Micromachines Corporation (Cambridge, MA, USA), to acquire high-resolution videos of the retina’s photoreceptors. Wavefront sensing was accomplished using a 780 nm superluminescent diode (Superlum, Cork, Ireland) and a 97-actuator deformable mirror (ALPAO, Montbonnot, France), which corrected higher-order aberrations in the wavefront of each subject’s eye. The AOSLO device has four imaging channels, though only one was used here: specifically, we used a 830nm laser diode to acquire confocal images of the retina at 15.7Hz (power at the eye: 200μW) and paired it a 15μm (0.77 Airy disk diameter) confocal pinhole.

Retinal stimulation was achieved using a 556 ±45nm fiber-coupled LED (MINT4F; ThorLabs Inc.) in Maxwellian view. In combination, all light used was less than a quarter of the maximum permissible exposure for the eye (ANSI Z136.1-2014).

### Imaging Protocol

After clearly explaining the study’s purpose and potential outcomes, participants provided written informed consent. The study eye was then dilated with 1% Tropicamide and 2.5% Phenylephrine. Control (non-stimulus) and stimulus trials were obtained from each study eye. Both control and stimulus acquisitions consisted of 10 seconds of recording, though stimulus acquisitions had a 7.1 μW, 62ms stimulus presented 2 seconds into the recording. All trials were initially collected using the 15 μm pinhole.

Each control trial was preceded by 15 minutes of dark adaptation, followed by ten acquisitions at a single location 8-10° temporal from each subject’s preferred retinal locus to maximize rod density and rod resolvability. Each stimulus trial always followed the control trial and was preceded by 10 minutes of dark adaptation, followed by ten acquisitions, with each preceded by 5 minutes of dark adaptation.

When all trials were collected, Apaeros software (Boston Micromachines) was used to register each video using strip-registration. All distortion-mitigated videos were co-registered and averaged, resulting in a super-average image for each participant.

### Intensity-based Optoretinography

To obtain intensity-based optoretinograms, we semi-automatically identified photoreceptor locations using semi-automated software (Mosaic; Translational Imaging Innovations; Hickory, NC). All cone and rod locations were marked by GY and verified by an experienced grader (RFC). Cones were identified and marked using the super-average image. Rod locations were determined using two methods (**Figure 1**): First, we identified rods in a similar manner to cones (**Figure 1A**). As rods are at the resolution limit of our device, even using a 15μm pinhole, these locations were determined using the super-average image *and* the average image of each video. Second, we extracted the location of all pixels that were *not* a part of cones (**Figure 1B**; hereafter called the ‘pixelwise’ rod iORG).

**Figure 1.**
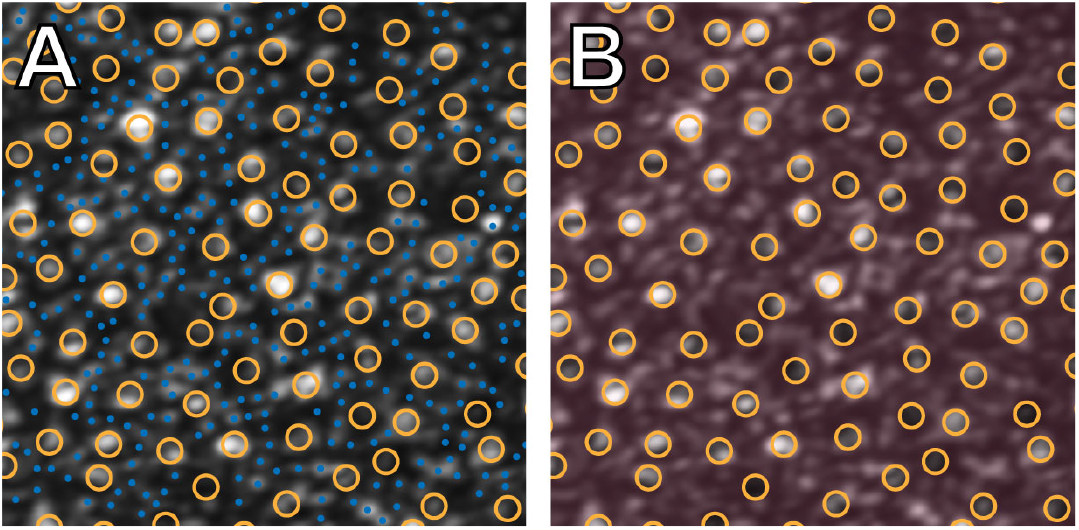
We determined the locations for analysis using two approaches. **A)** First, we obtained iORGs from cone (**orange circles**) and rod (**blue circles**) locations using semi-automated software. Regions with ambiguous or unclear cells were left unmarked. **B**) Second, we obtained iORGs from cone locations (**orange circles**) and all pixel locations that were not marked as cones (**red**).

Once all locations were determined, we used our custom ORG analysis software, *f*(Cell), to calculate the population root mean square (RMS) amplitude and half-amplitude implicit times for rod and cone locations. Briefly, the software operates as follows: Each frame in each video was standardized to a mean of 70 AU and a standard deviation of 35 AU to mitigate inter-video variability in mean and standard deviation. iORG signals were created from each cell in each video by averaging pixel intensities in a cylindrical column over time. We varied the cylinder radius as a function of cell size (cones=5px, rods=2px). Then, the pre-stimulus mean was subtracted from each iORG signal (**Figure 2B, C**).

**Figure 2:**
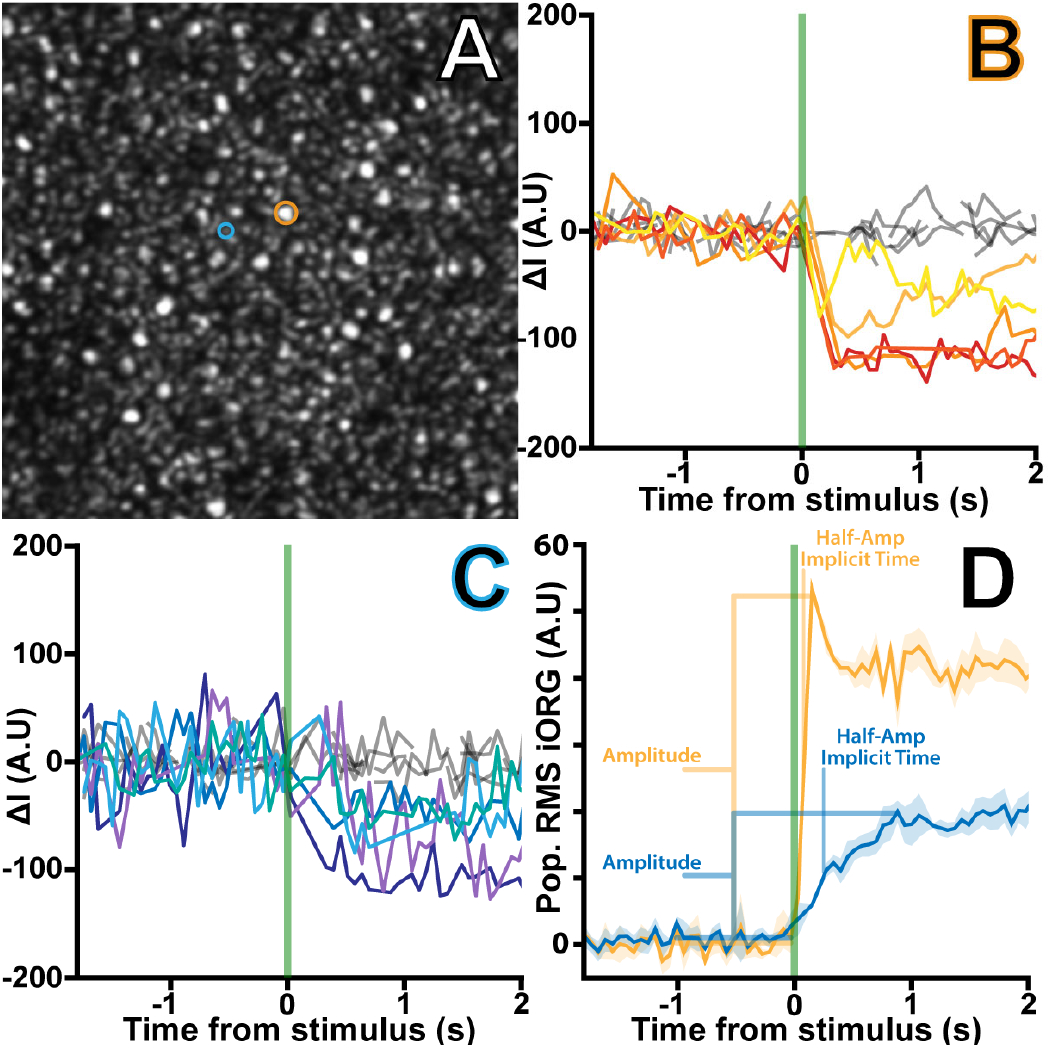
**A)** An example average image from a single video of the photoreceptor mosaic, with an example cone (**orange**) and rod (**blue**) highlighted. Across multiple acquisitions, the cone and rod highlighted in (**A**) exhibits intensity fluctuations in response to a stimulus (**B, cone**; **C, rod**). Control acquisitions are shown in gray. **D**) When these iORGs are summarized using RMS across either cones (**orange**) or rods (**blue**), these manifest as a rapid amplitude change in cones and a slow amplitude change in rods. Shaded regions are the RMS’ 95^th^ percentile confidence intervals.

We summarized all cone or rod iORGs in each video by obtaining the sample-wise RMS across all rod and cone locations (the ‘population RMS’). For each summary, average control RMS was subtracted from the stimulus RMS; all control-subtracted stimulus iORG RMS were then averaged (**Figure 2D**).

Finally, we extracted amplitude and half-amplitude implicit times from each video’s RMS (for intra-subject comparisons) and average iORG RMS waveform (for inter-subject comparisons) at each location. Amplitude was determined by subtracting the average pre-stimulus RMS from the 99th percentile of the post-stimulus RMS. We also extracted half-amplitude intrinsic time, defined as the time it takes to reach half the amplitude of the iORG RMS (**Figure 2D**). Differences between cone and rod metrics were evaluated using one-way ANOVAs; if significant, metric pairs were evaluated using Tukey’s Honest Significant Difference tests in SPSS (IBM, Armonk, NY). To evaluate relative differences between the cone and rod amplitudes independent of individual ORG variability, we also determined the ratio between the cone and rod amplitudes.

## Results

### The Rod Intensity-Based Optoretinogram

We successfully obtained rod and cone iORGs from an average of 249 cones (range: 157-336) and 972 rods (range: 654-1496) across all participants.

Across all participants, the average cone RMS amplitude (40.1 ±9.4) was higher and significantly different than the rod RMS amplitude (11.7 ±4.9; p<0.001, one-way ANOVA, Tukey HSD) derived from rod locations. Interestingly, rod RMS amplitude exhibited roughly half the inter-subject variability (9.4 vs 4.9).

On average, rod iORG RMS exhibited a slower response and a lower amplitude than cones. Rods and cones had average RMS half-amplitude implicit times of 270 ±71ms and 108 ±26ms, respectively (**Table 2**). Like amplitude, half-amplitude implicit times were significantly different between RMS derived from rod and cone locations (p=0.006, one-way ANOVA, Tukey HSD). We observed cone:rod amplitude ratios of 3.8±1.2.

**Table 2.** The average (± standard deviation) of metrics extracted from individual videos’ iORG RMS signals.

| Subject | Rod Locs Amplitude (A.U.) | Rod Pixelwise Amplitude (A.U.) | Cone Amplitude (A.U.) | Rod Locs Half-Amp Implicit Time (ms) | Rod Pixelwise Half-Amp Implicit Time (ms) | Cone Half-Amp Implicit time (ms) |
| --- | --- | --- | --- | --- | --- | --- |
| 36828 | 9.07 ± 2.1 | 8.7 ± 1.2 | 42.3 ± 2.9 | 298 ± 86 | 250 ± 88 | 75 ± 16 |
| 43556 | 8.9 ± 2.1 | 8.2 ± 1.5 | 37.9 ± 4.1 | 231 ± 47 | 195 ± 65 | 145 ± 15 |
| 86040 | 17.6 ± 3.1 | 14.0 ± 3.2 | 45.9 ± 4.9 | 278 ± 56 | 236 ± 30 | 149 ± 22 |
| 88735 | 13.2 ± 3.1 | 11.6 ± 3.3 | 25.2 ± 6.3 | 116 ± 98 | 98 ± 95 | 69 ± 40 |

### Pixelwise Rod Optoretinography

We successfully obtained pixelwise iORGs from rod-only locations, resulting in an average of 179,987 locations (range: 127,008-215,877) analyzed in each participant. Despite no segmentation and including considerably more sample points, we observed only minor differences between location-based and pixelwise rod iORG RMS signals within subjects (**Figure 3**). Further, there was no significant difference between metrics obtained using either method (**Figure 4, Table 2**; amplitude: p=0.98; half-amplitude implicit time: p=0.17; one-way ANOVA, Tukey HSD).

**Figure 3.**
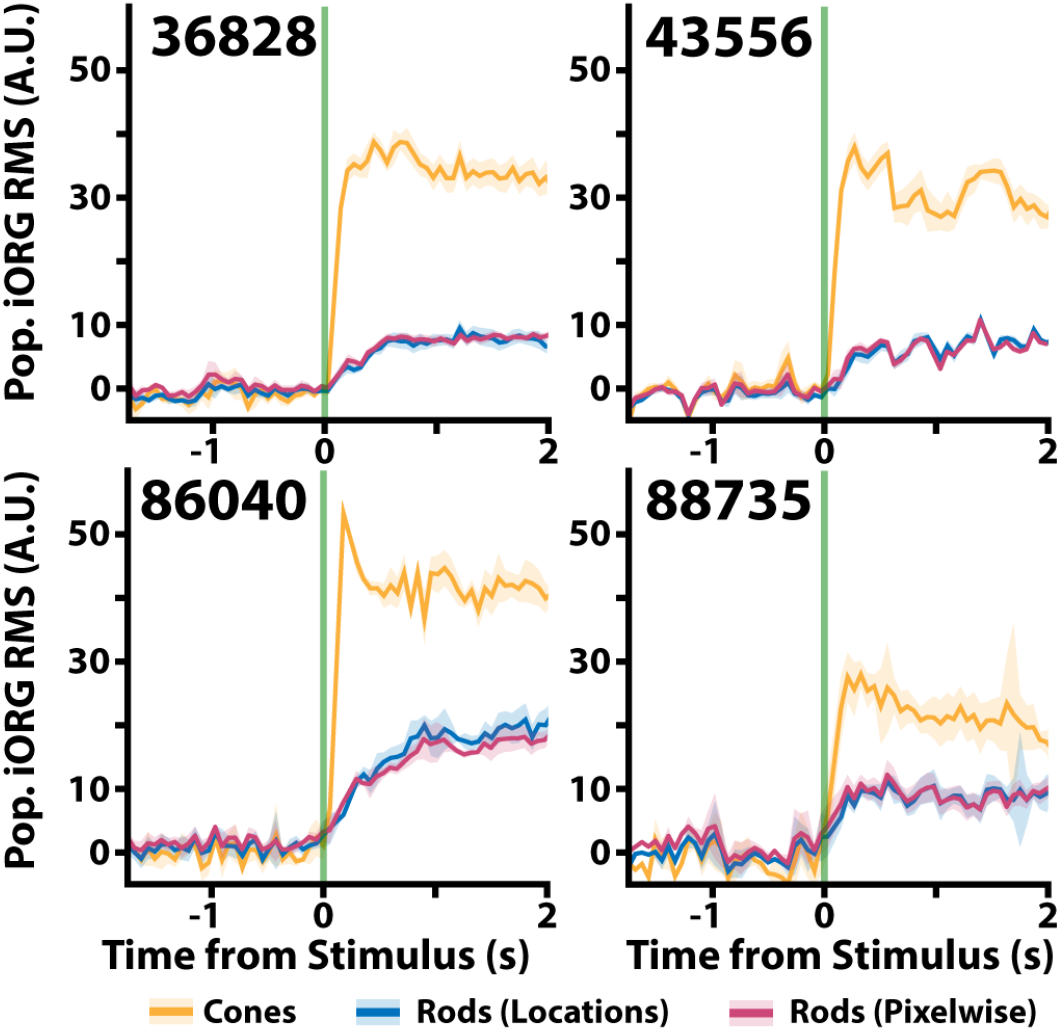
The control-subtracted cone and rod iORG RMS signals for each participant. The cone iORG RMS (**orange**) is variable across individuals, consistent with prior observations. The rod iORG RMS amplitude is more consistent across individuals. Notably, there is little difference between individual iORG RMS derived from identified rods (**blue**) vs iORG RMS derived from all pixels including rods (**red**).

**Figure 4.**
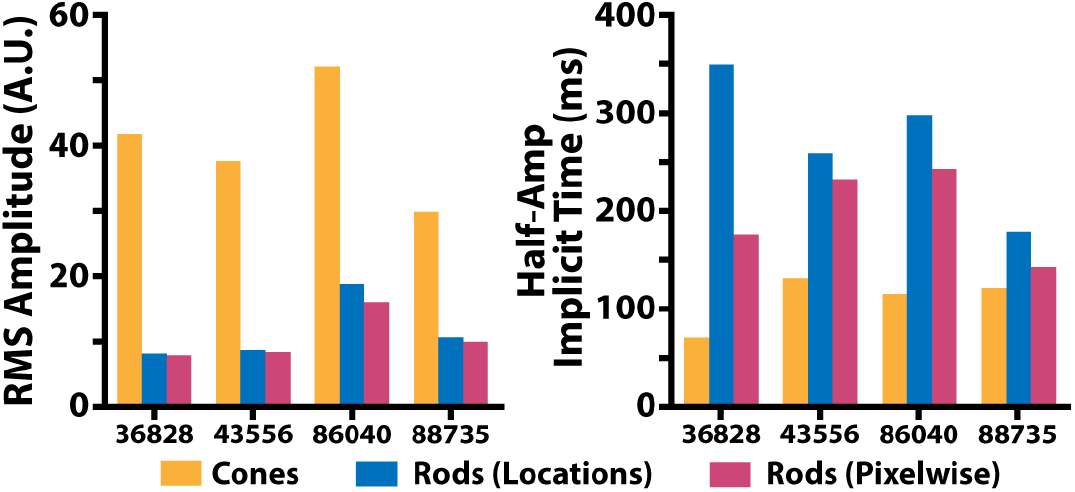
Each subjects’ average iORG RMS amplitude and implicit time followed similar patterns. (**left**) Cone RMS amplitudes had ratios always >2.5 the rod RMS amplitudes, and (**right**) cone half-amplitude implicit times always had ratios <0.7 the rod implicit times.

## Discussion

Electroretinography is considered the gold standard for evaluating photoreceptor function, but cannot resolve individual cells. Other functional tests, such as visual fields (VF), are widely used in the clinic but often prove unreliable. VF testing depends on consistent patient responses, which is not always feasible. Patients with motor impairments or those recovering from stroke, for example, may struggle to press the response button in time, creating the appearance of reduced retinal function even when the retina is intact. Optoretinography offers a way around these limitations by directly measuring photoreceptor activity. Because it bypasses patient response bias, it provides a more objective and reliable measure of retinal function, with potential for earlier disease detection and more accurate monitoring.

In this study, we report what we believe to be the first intensity-based optoretinograms of rod photoreceptors. Previous iORGs reports require laborious segmentation of individual photoreceptors, a process that is not practical for clinical use. Here, by applying a pixelwise approach to rod-only regions, we were able to evaluate photoreceptor function quickly and efficiently. Importantly, our results indicate that identifying individual rods is unnecessary, and that the rod RMS waveforms are nearly identical when obtained without individual rod identification (see **Figure 3**). This suggests that even when rods cannot be clearly resolved, their function can still be measured.

This pixelwise methodology has significant potential for real-world clinical applications. Rods are notoriously difficult to image, and eliminating the need for rod-level identification removes a major barrier. With this approach, rods can be assessed in a fast, scalable manner. Being able to directly assay rod photoreceptor function could be invaluable for monitoring disease progression and for evaluating the effectiveness of emerging rod-targeted therapies.

In addition to the benefits of this pixelwise methodology, we also identified several key findings: cone half-amplitude implicit times were consistently shorter than those of rods, cone RMS amplitudes were higher, and rod iORGs exhibited slower kinetics and lower amplitudes. These results were consistent with prior observations of rod optoretinograms using phase-based techniques.^43,54-56^ We also observed an average ratio of 3.8 between cone and rod iORG RMS amplitude; most of this variability originated from changes in the cone RMS. This highlights known differences in photoreceptor kinetics and reinforces the established distinction between cone-mediated photopic sensitivity and rod-mediated scotopic sensitivity.^57^ Importantly, the response patterns observed align with the known physiology of cones and rods, supporting the conclusion that the imaged structures were indeed rod photoreceptors rather than artifacts.

Notably, our human rod and cone iORG kinetics diverge from those of previous phase-based optoretinograms in rodent models.^53^ In contrast to earlier reports, human rods in our study had an implicit time within the 4-second measurement window, whereas rodent responses were markedly slower. These species-specific differences highlight the importance of direct human measurements for informing the design of next-generation functional imaging technologies.

Our study has several limitations. First, the dataset was derived from a subject pool composed exclusively of young (<50 years old), healthy individuals without known retinal pathology and with relatively low refractive corrections. Future work will investigate whether these findings hold in patients with rod degenerations. Second, the sample size was small (n = 4). We acknowledge that a larger cohort would increase the statistical power and generalizability of the results, and this will be an important direction for subsequent studies. Finally, the study did not include replication using alternative advanced optics scanning laser ophthalmoscopes (AOSLOs). Replicating these findings with other imaging systems will be critical for validating robustness and assessing broader applicability.

## Conclusion

More than 50 inherited retinal diseases are known, with rod photoreceptors serving as early indicators in nearly half of them. Given that current clinical imaging modalities cannot resolve the loss of individual rods, rod optoretinograms hold unique promise for transforming the early detection and precise monitoring of retinal disease. They provide a powerful means of detecting early functional changes in diseases marked by rod degeneration, such as Retinitis Pigmentosa, conditions like Age-related Macular Degeneration and certain forms of night blindness, where rod mosaics remain structurally intact, but physiologically impaired. Our imaging approach also allows us to distinguish between photoreceptor-driven and neurologically driven dysfunction, providing a pathway toward more targeted interventions.

In this work, we obtained intensity-based optoretinograms (iORGs) from rod photoreceptors using an adaptive optics scanning laser ophthalmoscope. We found that human rod iORG RMS waveforms have slower implicit times and lower amplitudes than cone iORG RMS waveforms. We also found that human rods reach implicit time faster than rodent models, highlighting the importance of studying human photoreceptor function directly to inform future technologies. Additionally, we determined that we can obtain rod iORG without counting individual rods, greatly simplifying their use, and making the approach more practical and scalable. With continued development, this method has potential to deliver a high-quality, accessible method for early detection of retinal diseases, thereby advancing the shared goal of preserving one of humanity’s most vital senses: vision.

## Data Availability

All data produced in the present study are available upon reasonable request to the authors.

## Acknowledgements

The authors would like to acknowledge the resources provided by the Ocular and Computer Vision Laboratory at Marquette University and the constructive feedback from colleagues. We would also like to thank Katie McKenney and Phyllis Summerfelt for patient recruitment, scheduling, and imaging. Finally, we are grateful to our families and friends for their encouragement.

## References

1. TD L. Photoreceptor physiology and evolution: cellular and molecular basis of rod and cone phototransduction - PubMed. The Journal of physiology. 2022 Nov;600(21)doi:10.1113/JP282058

2. Pearring JN, Salinas RY, Baker SA, Arshavsky VY. Protein sorting, targeting and trafficking in photoreceptor cells. Prog Retin Eye Res. Sep 2013;36:24–51. doi:10.1016/j.preteyeres.2013.03.002

3. Schneider N, Sundaresan Y, Gopalakrishnan P, et al. Inherited retinal diseases: Linking genes, disease-causing variants, and relevant therapeutic modalities. Prog Retin Eye Res. Jul 2022;89:101029. doi:10.1016/j.preteyeres.2021.101029

4. Dias MF, Joo K, Kemp JA, et al. Molecular genetics and emerging therapies for retinitis pigmentosa: Basic research and clinical perspectives. Prog Retin Eye Res. Mar 2018;63:107–131. doi:10.1016/j.preteyeres.2017.10.004

5. Curcio CA, Medeiros NE, Millican CL. Photoreceptor loss in age-related macular degeneration. Investigative Ophthalmology & Visual Science. 1996;37(7):1236–1249.

6. Hartong DT, Berson EL, Dryja TP. Retinitis pigmentosa. Lancet. Nov 18 2006;368(9549):1795–809. doi:10.1016/S0140-6736(06)69740-7

7. Hoffmann MB, Bach M, Kondo M, et al. ISCEV standard for clinical multifocal electroretinography (mfERG) (2021 update). Doc Ophthalmol. Feb 2021;142(1):5–16. doi:10.1007/s10633-020-09812-w

8. Carroll J, Kay DB, Scoles D, Dubra A, Lombardo M. Adaptive optics retinal imaging-clinical opportunities and challenges. Research Support, N.I.H., Extramural Research Support, Non-U.S. Gov’t Review. Current Eye Research. Jul 2013;38(7):709–721. doi:10.3109/02713683.2013.784792

9. Dubra A, Sulai Y, Norris JL, et al. Noninvasive imaging of the human rod photoreceptor mosaic using a confocal adaptive optics scanning ophthalmoscope. Biomedical Optics Express. Jul 1 2011;2(7):1864–1876. doi:10.1364/BOE.2.001864

10. Ahmed S, Son T, Ma G, Yao X. Polarization optical coherence tomography optoretinography: verifying light-induced photoreceptor outer segment shrinkage and subretinal space expansion. Neurophotonics. 2025;12(1):015005.

11. Chen S, Ni S, Jiménez-Villar A, Jian Y, Jia Y, Huang D. Optical coherence tomography split-spectrum amplitude-decorrelation optoretinography. Optics Letters. 2023;48(15):3921–3924.

12. Cooper RF, Brainard DH, Morgan JIW. Optoretinography of individual human cone photoreceptors. Optics Express. August 31 2020;28(26):39326–39339.

13. Gaffney M, Connor T, Cooper RF. Intensity-based optoretinography reveals subclinical deficits in cone function in retinitis pigmentosa. Frontiers in Ophthalmology. 2024;4

14. Gong Z, Shi Y, Liu J, Sabesan R, Wang RK. Light-adapted flicker-optoretinography based on raster-scan optical coherence tomography towards clinical translation. Biomedical Optics Express. 2024;15(10):6036–6051.

15. Jiang X, Liu T, Pandiyan VP, Slezak E, Sabesan R. Coarse-scale optoretinography (CoORG) with extended field-of-view for normative characterization. Biomedical Optics Express. 2022;13(11):5989–6002.

16. Jiang X, Liu T, Pandiyan VP, Slezak E, Sabesan R. Coarse-scale optoretinography (CoORG) with extended field-of-view for normative characterization Biomedical Optics Express. 2022;13(11):5989–6002.

17. Kim T-H, Ding J, Yao X. Intrinsic signal optoretinography of dark adaptation kinetics. Scientific Reports. 2022;12

18. Lassoued A, Zhang F, Kurokawa K, et al. Cone photoreceptor dysfunction in retinitis pigmentosa revealed by optoretinography. Proceedings National Academy of Sciences, USA. 2021;118(47):e2107444118. doi:10.1073/pnas.2107444118

19. Ma G, Son T, Kim TH, Yao X. Functional optoretinography: concurrent OCT monitoring of intrinsic signal amplitude and phase dynamics in human photoreceptors. Biomed Opt Express. May 1 2021;12(5):2661–2669. doi:10.1364/BOE.423733

20. Pandiyan VP, Jiang X, Maloney-Bertelli A, Kuchenbecker JA, Sharma U, Sabesan R. High-speed adaptive optics line-scan OCT for cellular-resolution optoretinography. Biomedical Optics Express. 2020;11(9):5274–5296.

21. Pandiyan VP, Schleufer S, Slezak E, et al. Characterizing cone spectral classification by optoretinography. Biomedical Optics Express. 2022;13(12):6574–6594.

22. Roorda A. Optoretinography is coming of age. Proceedings National Academy of Sciences, USA. 2021;118(51):e2119737118. doi:10.1073/pnas.2119737118

23. Siddiqui A, Gaffney M, Cooper RF. Assessing the feasibility and repeatability of light-adapted intensity-based optoretinography. Investigative Ophthalmology & Visual Science. 2023;64(8):3347.

24. Tan B, Li H, Zhuo Y, et al. Light-evoked deformations in rod photoreceptors, pigment epithelium and subretinal space revealed by prolonged and multilayered optoretinography. Nature Communications. 2024;15:5156.

25. Tomczewski S, Węgrzyn P, Wojtkowski M, Curatolo A. Chirped flicker optoretinography for in vivo characterization of human photoreceptors’ frequency response to light. Optics Letters. 2024;

26. Vienola KV, Valente D, Zawadzki RJ, Jonnal RS. Velocity-based optoretinography for clinical applications. Optica. 2022;9(10):1100–1108.

27. Wendel BJ, Pandiyan VP, Liu T, et al. Multimodal high-resolution imaging in retinitis pigmentosa: A comparison between optoretinography, cone density, and visual sensitivity. Investigative Ophthalmology & Visual Science. 2024;65(10):45.

28. Wongchaisuwat N, Amato A, Yang P, et al. Optical coherence tomography split-spectrum amplitude-decorrelation optoretinography detects early central cone photoreceptor dysfunction in retinal dystrophies. Translational Vision Science & Technology. 2024;13(10):5.

29. Xu P, Cooper RF, Jiang YY, Morgan JIW. Parafoveal cone function in choroideremia assessed with adaptive optics optoretinography. Scientific Reports. 2024;14(1):8339.

30. Jonnal RS, Rha J, Zhang Y, Cense B, Gao W, Miller DT. In vivo functional imaging of human cone photoreceptors. Optics Express. 2007;14(24):16141–16160.

31. Cooper RF, Tuten WS, Dubra A, Brainard DH, Morgan JIW. Non-invasive assessment of human cone photoreceptor function. Biomedical Optics Express. 2017;8(11):5098–5112.

32. Rha J, Schroeder B, Godara P, Carroll J. Variable optical activation of human cone photoreceptors visualized using short coherence light source. Optics Letters. 2009;34(24):3782–3784.

33. Grieve K, Roorda A. Intrinsic signals from human cone photoreceptors. Investigative ophthalmology & visual science. Feb 2008;49(2):713–719. doi:49/2/713 [pii] 10.1167/iovs.07-0837

34. Yao X, Kim T. Fast intrinsic optical signal correlates with activation phase of phototransduction in retinal photoreceptors. Experimental Biology and Medicine. 2020;245(13):1087–1095.

35. Warner RL, Brainard DH, Morgan IWM. Repeatability and reciprocity of the cone optoretinogram. Biomedical Optics Express. 2022;13(12):6561–6573.

36. Bedggood P, Metha A. Optical imaging of human cone photoreceptors directly following the capture of light. PLoS One. 2013;8(11):e79251.

37. Messner A, Dos Santos VA, Puchner S, et al. The impact of photopigment bleaching on the human rod photoreceptor subretinal space measured via optical coherence tomography. Investigative ophthalmology & visual science. 2024;65(3):20.

38. Zhang Q, Lu R, Curcio CA, Yao X. In vivo confocal intrinsic optical signal identification of localized retinal dysfunction. Investigative ophthalmology & visual science. 2012;53(13):8139–8145.

39. Bedggood P, Metha A. Variability in bleach kinetics and amount of photopigment between individual foveal cones. Investigative ophthalmology & visual science. 2012;53(7):3673–3681.

40. Hillmann D, Spahr H, Pfäffle C, Sudkamp H, Franke G, Hüttmann G. In vivo optical imaging of physiological responses to photostimulation in human photoreceptors. Proceedings of the National Academy of Sciences of the United States of America. 2016;113(46):13138–13143.

41. Pfäffle C, Spahr H, Kutzner L, et al. Simultaneous functional imaging of neuronal and photoreceptor layers in living human retina Optics Letters. 2019;44(23):5671–5674.

42. Zhang F, Kurokawa K, Lassoued A, Crowell JA, Miller DT. Cone photoreceptor classification in the living human eye from photostimuluation-induced phase dynamics. Proceedings of the National Academy of Sciences of the United States of America. 2019;116(16):7951–7956.

43. Azimipour M, Valente D, Vienola KV, Werner JS, Zawadzki RJ, Jonnal R. Optoretinogram: optical measurement of human cone and rod photoreceptor responses to light. Optics Letters. 2020;45(17):4658–4661. doi:10.1364/OL.398868

44. Pandiyan VP, Bertelli AM, Kuchenbecker JA, et al. The optoretinogram reveals the primary steps of phototransduction in the living human eye. Science Advances. 2020;6(37)doi:10.1126/sciadv.abc1124

45. Yao XC, Yamauchi A, Perry B. Rapid optical coherence tomography and recording functional scattering changes from activated frog retina. Applied Optics. 2005;44(11):2019–2023.

46. Tomczewski S, Węgrzyn P, Borycki D, Auksorius E, Wojtkowski M, Curatolo A. Light-adapted flicker optoretinograms captured with a spatio-temporal optical coherence-tomography (STOC-T) system. Biomedical Optics Express. 2022;13(4):2186–2201.

47. Pfäffle C, Puyo L, Spahr H, Hillmann D, Miura Y, Hüttmann G. Unraveling the functional signals of rods and cones in the human retina: separation and analysis. Frontiers in Ophthalmology. 2024;4

48. Pfäffle C, Spahr H, Gercke K, et al. Phase-sensitive measurements of depth-dependent signal transduction in the inner plexiform layer. Frontiers in Medicine. 2022;9

49. Zhang F, Kurokawa K, Bernucci MT, et al. Revealing how color vision phenotype and genotype manifest in individual cone cells. Investigative ophthalmology & visual science. 2021;62(2):8.

50. Pandiyan VP, Nguyen PT, Pugh Jr. EN, Sabesan R. Human cone elongation responses can be explained by photoactivated cone opsin and membrane swelling and osmotic response to phosphate produced by RGS9-catalyzed GTPase. Proceedings National Academy of Sciences, USA. 2022;119(39):e2202485119.

51. Curcio CA, Sloan KR, Kalina RE, Hendrickson AE. Human photoreceptor topography. J Comp Neurol. Feb 22 1990;292(4):497–523. doi:10.1002/cne.902920402

52. Lee SCS, Martin PR, Grunert U. Topography of Neurons in the Rod Pathway of Human Retina. Invest Ophthalmol Vis Sci. Jul 1 2019;60(8):2848–2859. doi:10.1167/iovs.19-27217

53. Zhang P, Zawadzki RJ, Goswami MN P.T.,, Yarov-Yarovoy V, Burns ME, Pugh EN, Jr. In vivo optophysiology reveals that G-protein activation triggers osmotic swelling and increased light scattering of rod photoreceptors. Proceedings of the National Academy of Sciences of the United States of America. 2017;114(14):E2937–E2946.

54. Schnapf JL, Copenhagen DR. Differences in the kinetics of rod and cone synaptic transmission. Nature. 1982;296(5860):862–864. doi:10.1038/296862a0

55. Thoreson WB. Kinetics of synaptic transmission at ribbon synapses of rods and cones. Mol Neurobiol. 2007;36(3):205–223. doi:10.1007/s12035-007-0019-9

56. Lassoued A, Zhang F, Kurokawa K, et al. Cone photoreceptor dysfunction in retinitis pigmentosa revealed by optoretinography. Proc Natl Acad Sci U S A. 2021;118(47):e2107444118. doi:10.1073/pnas.2107444118

57. Fu Y. Phototransduction in rods and cones. In: Kolb H, Fernandez E, Jones B, et al, eds. Webvision: The Organization of the Retina and Visual System [Internet]. Salt Lake City, UT: University of Utah Health Sciences Science; 1995 -. Updated July 30, 2018. Accessed August 29, 2025. https://www.ncbi.nlm.nih.gov/books/NBK52768/

